# Dynamic data-driven meta-analysis for prioritisation of host genes implicated in COVID-19

**DOI:** 10.1101/2020.08.27.20182238

**Authors:** Nicholas Parkinson, Natasha Rodgers, Max Head Fourman, Bo Wang, Marie Zechner, Maaike C. Swets, Jonathan E. Millar, Andy Law, Clark D. Russell, J. Kenneth Baillie, Sara Clohisey

## Abstract

The increasing body of literature describing the role of host factors in COVID-19 pathogenesis demonstrates the need to combine diverse, multi-omic data to evaluate and substantiate the most robust evidence and inform development of therapies.

Here we present a dynamic ranking of host genes implicated in human betacoronavirus infection (SARS-CoV-2, SARS-CoV, MERS-CoV, seasonal coronaviruses). Researchers can search and review the ranked genes and the contribution of different experimental methods to gene rank at https://baillielab.net/maic/covid19.

We conducted an extensive systematic review of experiments identifying potential host factors. Gene lists from diverse sources were integrated using Meta-Analysis by Information Content (MAIC). This previously described algorithm uses data-driven gene list weightings to produce a comprehensive ranked list of implicated host genes.

From 32 datasets, the top ranked gene was PPIA, encoding cyclophilin A, a drug-gable target using cyclosporine.Other highly-ranked genes included proposed prognostic factors (*CXCL10, CD4, CD3E*) and investigational therapeutic targets (*IL1A*) for COVID-19. Gene rankings also inform the interpretation of COVID-19 GWAS results, implicating *FYCO1* over other nearby genes in a disease-associated locus on chromosome 3.

As new data are published we will regularly update list of genes as a resource to inform and prioritise future studies.

## Introduction

There are multiple sources of information that associate host genes with SARS-CoV-2 viral replication, the subsequent host immune response and the ensuing pathophysiology. Integrating these sources of information may provide more robust evidence associating specific genes and proteins with key processes underlying the mechanisms of disease. This is needed in order to make informed judgements about new therapies for inclusion in model studies and clinical trials.

The pace of new research into COVID-19 pathophysiology, including host dependency factors, immune responses, and genetics, has made it nearly impossible to read every report. In addition, assessing the quality and relevance of new evidence is difficult, time-consuming, and requires a high level of expertise. Information from diverse sources has varying quality, scale, and relevance to host responses to SARS-CoV-2. Computational approaches can aid data evaluation and integration. Simple, intuitive methods have a conceptual advantage for translation to decision-making: if both the processes and results are easily comprehensible, then it is easier for human users to trust the conclusions.

SARS-CoV-2 is a betacoronavirus with a 30kb single-stranded positive-sense RNA genome, and is genetically similar to other human coronaviruses: SARS-CoV, MERS-CoV and the seasonal ‘common cold’ 229E, OC43, HKU1 and NL63 coronaviruses. Like all viruses, SARS-CoV-2 relies on host machinery to replicate. Host dependency factors represent an attractive target for new therapeutics, as evolution of drug resistance is expected to be slower for host-directed than viral-directed therapies.^1^

Treatment directly targeting viral replication can target viral proteins (e.g. remdesivir^2^), or host proteins upon which the virus depends.^3^ Host-targeted therapies may have an important role in infectious diseases in general, and the only treatment so far found to reduce mortality in COVID-19 - dexamethasone^4^ - is likely to act by targeting host immune-mediated organ damage.^5^ Other host-directed treatments (e.g. anakinra, tocilizumab, sarilumab, mavrilimumab), repurposed from other indications, are currently under investigation.^2,6,7,8,9,10^

In this analysis we systematically identify and combine existing data from human betacoronavirus research to generate a comprehensive ranked list of host genes as a resource to inform further work on COVID-19.

To identify existing literature which could provide informative datasets for host gene prioritisation, we conducted a systematic review of published studies and pre-print manuscripts pertaining to host gene involvement in human betacoronavirus infection and associated disease. Results from identified studies, in the form of lists of implicated host factor genes, were combined using meta-analysis by information content (MAIC),^3^ an approach we previously developed to identify host genes necessary for Influenza A virus (IAV) replication. We have previously demonstrated that the MAIC algorithm successfully predicts new experimental results from an unseen future experiment.^3^ Our gene prioritisation results both recapitulate existing understanding of COVID-19 pathophysiology and highlight key host factors and potential therapeutic targets that have to date been largely overlooked.

## Methods

### Meta-analysis by information content (MAIC)

MAIC allows the combination of data from diverse sources without prior assumptions regarding the quality of each individual data source. The MAIC approach begins with the following assumptions:

1. There exists a set of true positives: host genes involved in COVID-19 pathogenesis.
2. A gene is more likely to be a true positive if it is found in multiple experiments.
3. A gene is more likely to be a true positive if it occurs in a list containing a higher proportion of genes with supporting evidence from multiple sources.
4. Due to experimental biases, the evidence that a gene is a true positive is further increased if it is found across experimental types.

With these assumptions, MAIC allows the quantification of the information content in a gene list by comparing that list to the results from other experiments that might reasonably be expected to find some of the same genes. Input gene lists can be categorised by data type (tbl. 2), allowing comparison both within and between methodologies. MAIC produces a weighting factor for each experiment, and this weighting is used to calculate a score for each gene. The analysis then produces a final ranked list of genes based on this score, which summarises the combined evidence from all input sources of that particular gene being involved in SARS-CoV-2 pathogen-host interaction. A full description of the MAIC algorithm can be found in our original report.^3^

### Data eligibility

Inclusion and exclusion criteria are shown in tbl. 1. To complement emerging data pertaining to the novel SARS-CoV-2, we included studies of other human coronaviruses. Included methodologies are shown in tbl. 2.

**Table 1:** Entry Criteria

| Inclusion | Exclusion |
| --- | --- |
| Virus studied: SARS-CoV, SARS-CoV-2, MERS-CoV, HCoV-229E, HCoV-HOC43, HCoV-HKU1, HCoV-NL63<br>Methodology present in tbl. 2 | Candidate in vitro or in vivo gene, transcript or protein studies and screens - defined here as <50 genes, transcripts or proteins investigated<br>Candidate-gene human genetic studies<br>< 5 hosts in virus group or control group in patient studies |
|  | Meta-analyses, in silico analyses, re-analysis of data published elsewhere |

**Table 2:** Methodologies accepted for inclusion in meta-analysis and associated labels

| Accepted Methodologies | MAIC category |
| --- | --- |
| CRISPR screen | CRISPR SCREEN |
| RNAi screen | RNAI |
| Protein-protein interaction e.g. yeast-2-hybrid screen | PROTEIN-PROTEIN INTERACTION |
| Host proteins incorporated into virion or virus like particle | VIRUS |
| Genetic Association Studies | HUMAN GENETICS, NON-HUMAN GENETICS |
| Transcriptomics eg RNA-Seq, scRNA-Seq | TRANSCRIPTOMICS |
| Proteomic studies eg. mass-spectrometry | PROTEOMICS |
| Selected gene set screens | GENE SET SCREEN |

### Literature search

A systematic literature search of PubMed was conducted on 28/04/2020 and updated weekly until 06/07/2020. We used the following search strategy, with no date or language restrictions: > Keywords: (“Coronavirus” OR “Severe Acute Respiratory Syndrome” OR “Middle East Respiratory Syndrome” OR “Sars-CoV-2” OR “COVID-19”) AND ((gene*.[Title/Abstract]) OR (genom*[Title/Abstract]) OR (transcript*[Title/Abstract]) OR (protein*[Title/Abstract]) OR (“Susceptibility”[Title/Abstract]) OR (siRNA[All Fields]))

Potentially relevant pre-print manuscripts were identified by screening all papers categorised as COVID-19-related in the bioRxiv and medRxiv servers. Titles and abstracts of all returned papers were first assessed for relevance and duplication by a single member of the review team. Following this, full-length texts were obtained and an in-depth review was carried out by two further reviewers, independently, in order to confirm eligibility according to tbl. 2 and tbl. 1. In cases where a consensus was not reached, a third reviewer appraised the paper. This method ensured each paper was assessed for eligibility by a minimum of three independent reviewers. Relevant data, as shown in tbl. 3, was extracted from each reviewed paper.

**Table 3:**
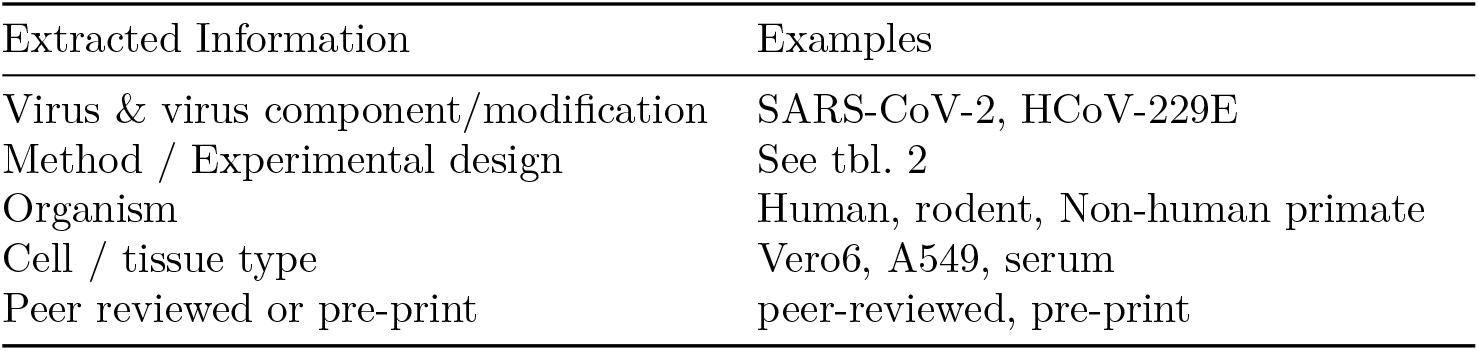
Data extracted from each publication

| Extracted Information | Examples |
| --- | --- |
| Virus & virus component/modification | SARS-CoV-2, HCoV-229E |
| Method / Experimental design | See tbl. 2 |
| Organism | Human, rodent, Non-human primate |
| Cell / tissue type | Vero6, A549, serum |
| Peer reviewed or pre-print | peer-reviewed, pre-print |

### Gene list extraction and categorisation

Relevant gene lists were identified and extracted. Datasets were excluded from the analysis where insufficient data were available to construct a meaningful unbiased gene list, for example where results for only a non-systematically selected subset of genes of interest were reported. Gene lists were categorised based on methodology as shown in tbl. 2.

Gene list rankings were preserved where possible, if sufficient numerical data were available. Rankings were based on significance or magnitude of effect. Adjusted measures of significance, usually adjusted *p*, were prioritised over raw *p* and *logFC* to determine ranking where multiple values were available. For studies reporting comparisons at multiple time points, genes were ranked based on the minimum *p* across all comparisons. To exclude irrelevant genes, a significance or effect size threshold was applied to all lists. This was either the threshold used by the authors for reporting, or where full data were provided this was determined as adjusted *p* < 0.05, |*z score*| > 1.96 or |*logFC*| > 1.5 depending on available values.

Gene, transcript and protein names or identification numbers were converted to the associated HGNC gene symbol, or an equivalent Ensembl or Refseq symbol where no HGNC symbol existed. Non-primate genes were mapped to their human homologues using the NCBI Homologene database,^11^ or excluded from the analysis if no human homologue could be identified.

### Gene set enrichment analysis

Rank-based gene set enrichment analysis was performed using the package FGSEA in R version 3.5.2, with genes ranked by MAIC score.^12^ p-values were estimated using an empirical probability distribution based on 10^6^ permutations. Gene set over-representation in the top 100 genes was analysed by a Fisher’s exact test as implemented in Enrichr.^13^ The Benjamini-Hochberg procedure was used to control the false discovery rate (*FDR <* 0.05) for both methods.

## Results

### Systematic review of the literature

We identified a total of 31 studies with available data meeting our eligibility criteria (12 pre-print manuscripts and 19 peer-reviewed studies), yielding 32 gene lists (Supplementary Figure 1, Supplementary Table 1). The included gene lists comprised 11 ranked and 21 unranked lists, in 8 experimental categories, with list lengths ranging from three to 9,967 genes (median 61). The datasets included 3 genetic perturbation screens (CRISPR, RNAi and interferon-stimulated gene overexpression), 3 genetic studies (of which 2 were in humans), 7 protein-protein interaction studies, 7 proteomic and 12 transcriptomic studies.

### MAIC analysis of identified studies

Of 5,418 genes implicated in human betacoronavirus infection in these 32 datasets, 4,150 are supported by a single paper only, 629 had evidence from more than one source within the same experimental category, and 639 are supported by data from multiple study types. Although extensive within-category overlap was seen for transcriptomic studies, there was less concordance within categories such as proteomics and protein-protein interaction. As with our previous study of influenza, contributions from one CRISPR screen dominate the overall information content (fig. 1 B). This was due in part to list length and hence contribution to scores for multiple lower-ranking genes. Information contributions for the top 100 genes are more balanced (fig. 1 C). The MAIC score distribution (fig. 1 D) reflects the degree of cross-category overlap, with the highest ranked genes supported by data from three distinct experimental categories.

**Figure 1:**
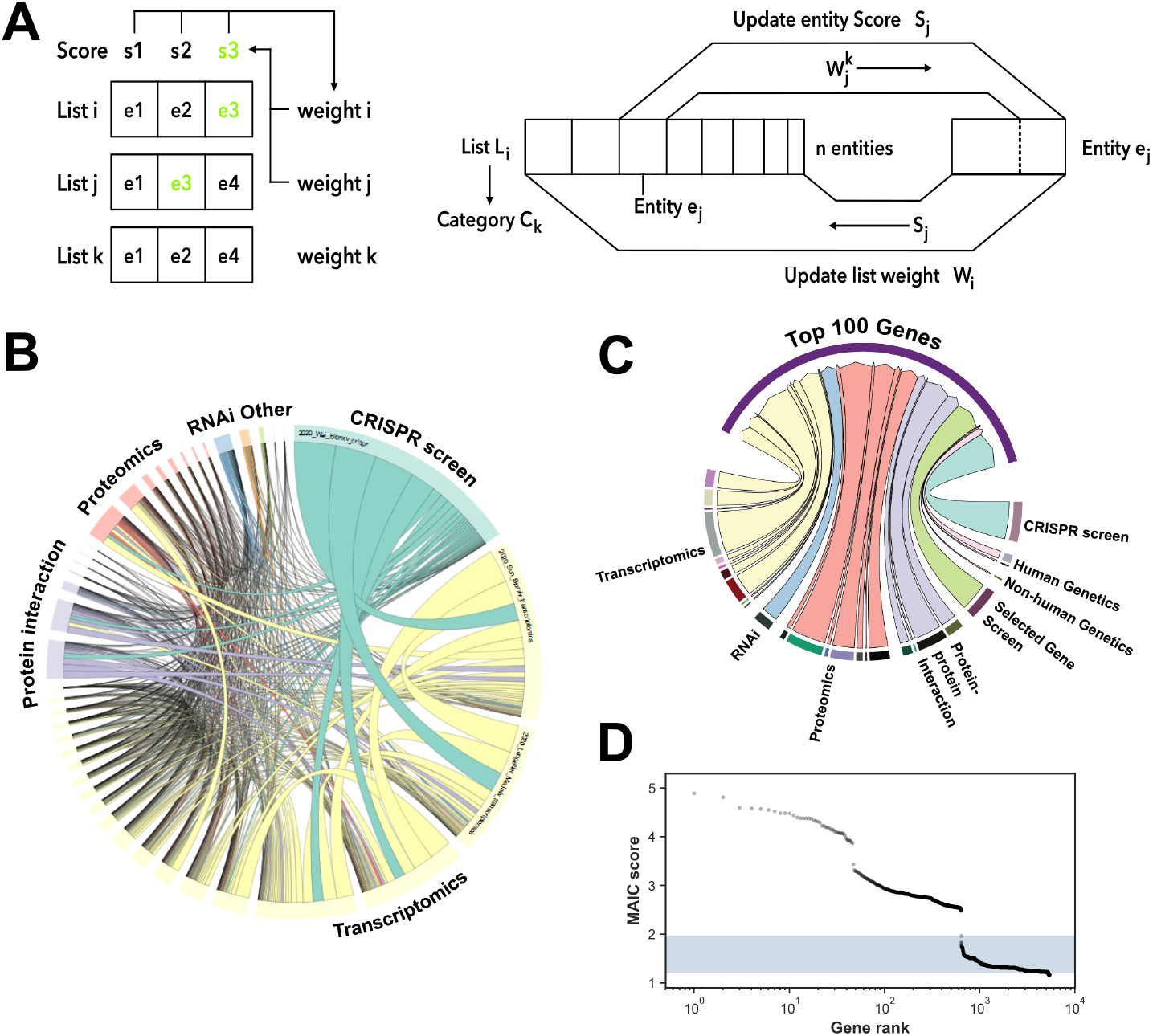
Overview of MAIC approach. (A) Schematic showing the operation of MAIC. Each entity in a list is given a score, based on overlap with other lists and rank where relevant, and each list is given a weight determined by the scores of its constituent entities. Entity scores are iteratively updated using list weights, and list weights are updated using entity scores, until convergence occurs. (B) Circular plot showing overlap between different data sources included in MAIC. Size of data source blocks is proportional to the summed information content (MAIC scores) of the input list. Lines are coloured according to the dominant data source. Data source categories share the same colour; the largest categories and data sources are labelled (see supplementary information for full source data). (C) Relative information contributions (determined by sum of MAIC score contributions) of each experimental category to the evidence base for the top 100 genes in the MAIC output. (D) Distribution of MAIC scores by gene rank. The shaded region indicates the range of possible scores for a gene supported by a single gene list only. Beyond ranks around 700 in this study, gene scores approach baseline, indicating they have little corroborative evidence.

**Figure 2:**
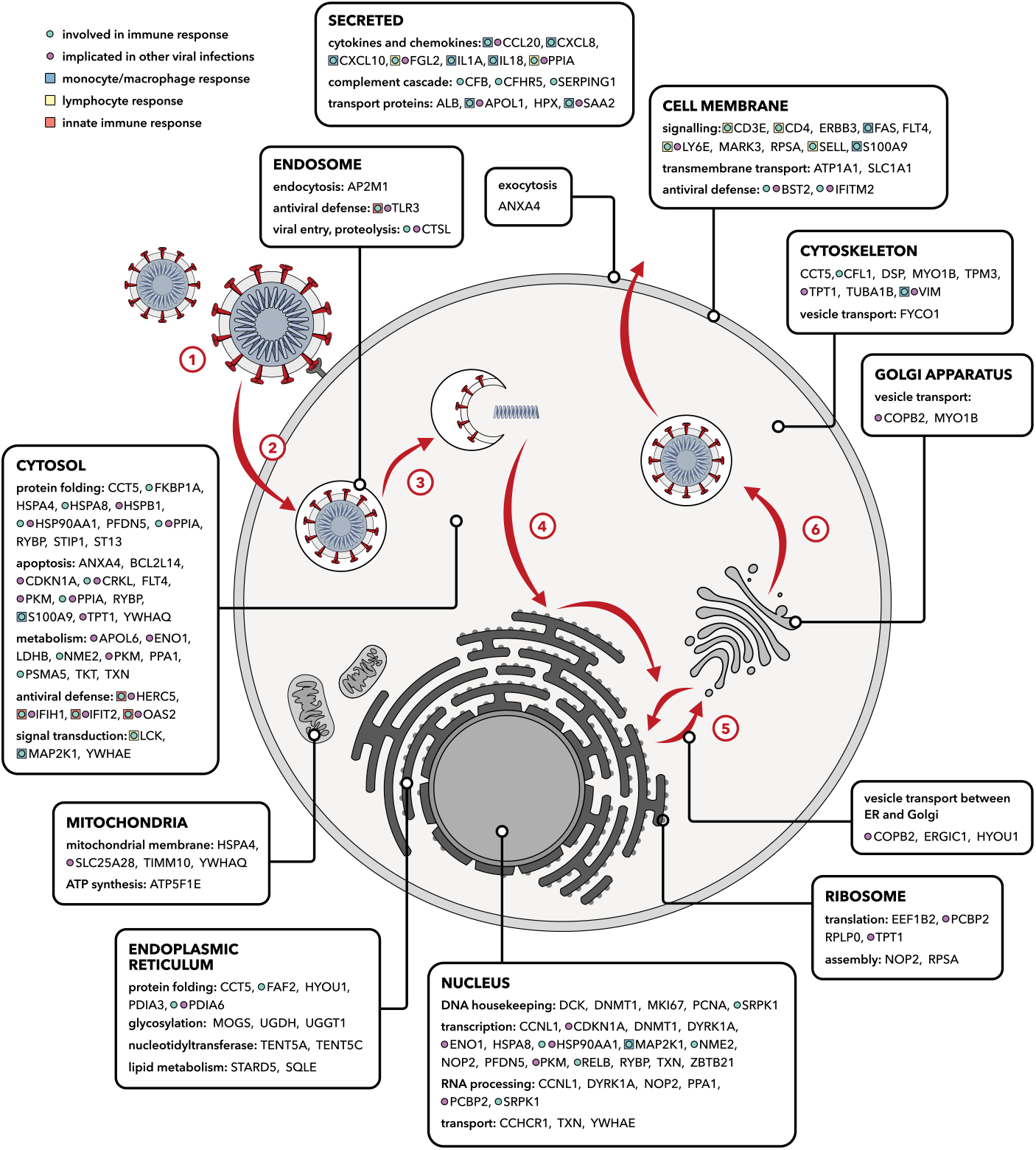
Cellular functions of the 100 highest ranked genes in the MAIC output. Protein products of these genes have diverse cellular locations and are associated with numerous processes relevant to the viral life cycle and host immune system. Stages of the betacoronavirus life cycle: (1) S protein-mediated attachment to the cell surface. (2) Endocytosis. (3) Membrane fusion and viral genome release into the cytoplasm. (4) Assembly of the replication-transcription complex, translation of mRNA. (5) Viral replication and virion assembly. (6) Virion maturation, budding and translocation of vesicles.

The highest ranking genes and their contributing evidence sources are shown in fig. 3 A and Supplementary Table 2. Top genes include *IL1A*^14^ and other components of the innate and adaptive immune systems (such as *CXCL10, CD4* and *TLR3*), which have previously been shown to contribute to COVID-19 pathogenesis. Other top genes have not previously received much attention in the context of coronavirus infection. These include *PPIA* (cyclophilin A), and *RYBP* (RING1 and YY1 binding protein), which play roles in protein folding, transcriptional repression, regulation of proteasomal degradation and apoptosis.

**Figure 3:**
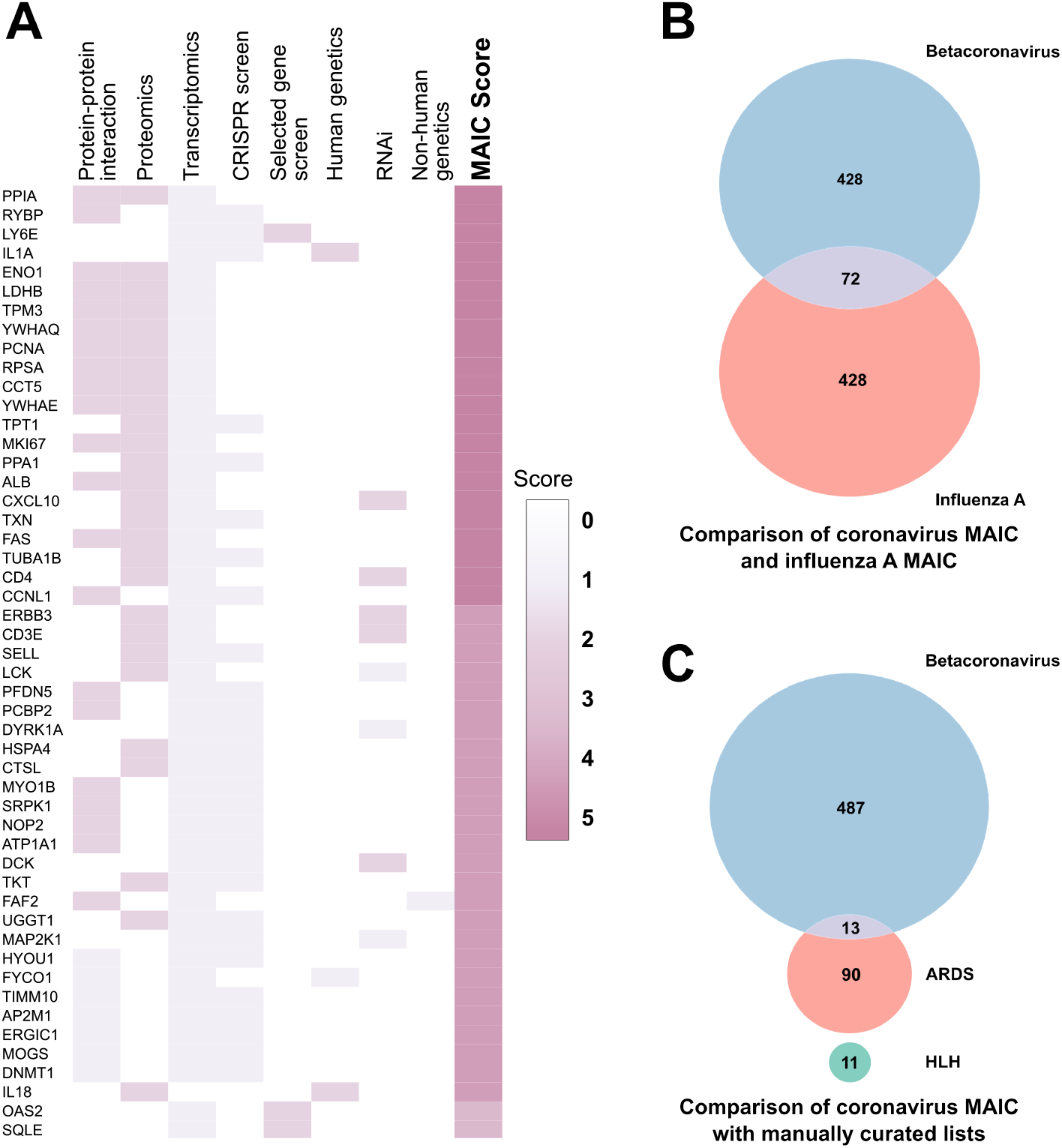
Highest ranked genes in the MAIC output and overlap with other conditions. (A) Heatmap of the top 50 genes implicated in SARS-CoV-2 infection, as ranked by the MAIC algorithm. The heatmap shows the information sources contributing to each of the top genes, by experimental category. Full details of all scored genes, including specific studies contributing to each, are given in Supplementary Table 2. (B) Venn diagram of overlap between the top 500 hits from this study and the top 500 hits from our previous MAIC analysis of Influenza A virus. (C) Venn diagram of overlap between the top 500 hits from this study and manually curated lists from available literature on HLH and ARDS.

An up-to-date prioritised list of implicated genes is available at bail-lielab.net/maic/covid19. We will repeat the analysis regularly as new data become available.

### Overlap with genes involved in influenza A, ARDS and familial HLH pathogenesis

Death in severe COVID-19 is usually a consequence of lung injury leading to ARDS (Acute Respiratory Distress Syndrome), a final common pathway that can occur in any severe acute respiratory infection. Host susceptibility factors in COVID-19 may be shared with other infections or ARDS. We compared the output from our analysis to our previous MAIC analysis of Influenza A virus.^3^ Among the top 500 ranked genes from each output, we found 72 overlapping genes (fig. 3 B), including a large number of RNA-binding, ribosome-associated and chaperone genes. Unexpectedly few immune related genes overlapped. This is surprising as both viruses are single-stranded RNA viruses and despite differing in sense, intracellular pathogen detection mechanisms are expected to be similar.

To expand this analysis we manually curated genes associated with ARDS and determined the overlap with our MAIC output (Supplementary Table 4). Among the top 500 ranked genes from the coronavirus MAIC output we found an overlap of 13 genes with the ARDS list (consisting of 103 genes) (fig. 3 C). Here we saw a number of genes associated with innate immunity and modulation of inflammation (*TNFα, IL6, IL18, CCL2, IL1B, TLR1, IL13, NFκBIA*).

The inflammatory profile, including hyperferritinaemia, observed in COVID-19 has led to the suggestion that a form of secondary haemophagocytic lympho-histiocytosis (HLH), a hyper-inflammatory syndrome, could be occurring.^15^ We manually curated genes involved in the familial form of this syndrome and compared these with our MAIC output, finding no overlap, although only eleven genes were found in the literature to be associated with familial HLH.

### Pathway analysis of MAIC output

To better understand the biological functions of the most strongly implicated genes, we performed gene set enrichment analysis in ten databases of functional annotations. We used two complementary methods, assessing enrichment either in terms of rank distribution across the whole dataset (permissive) or in over-representation in the top 100 genes only (conservative). There was extensive overlap between these approaches (fig. 4 A and Supplementary Table 3).

**Figure 4:**
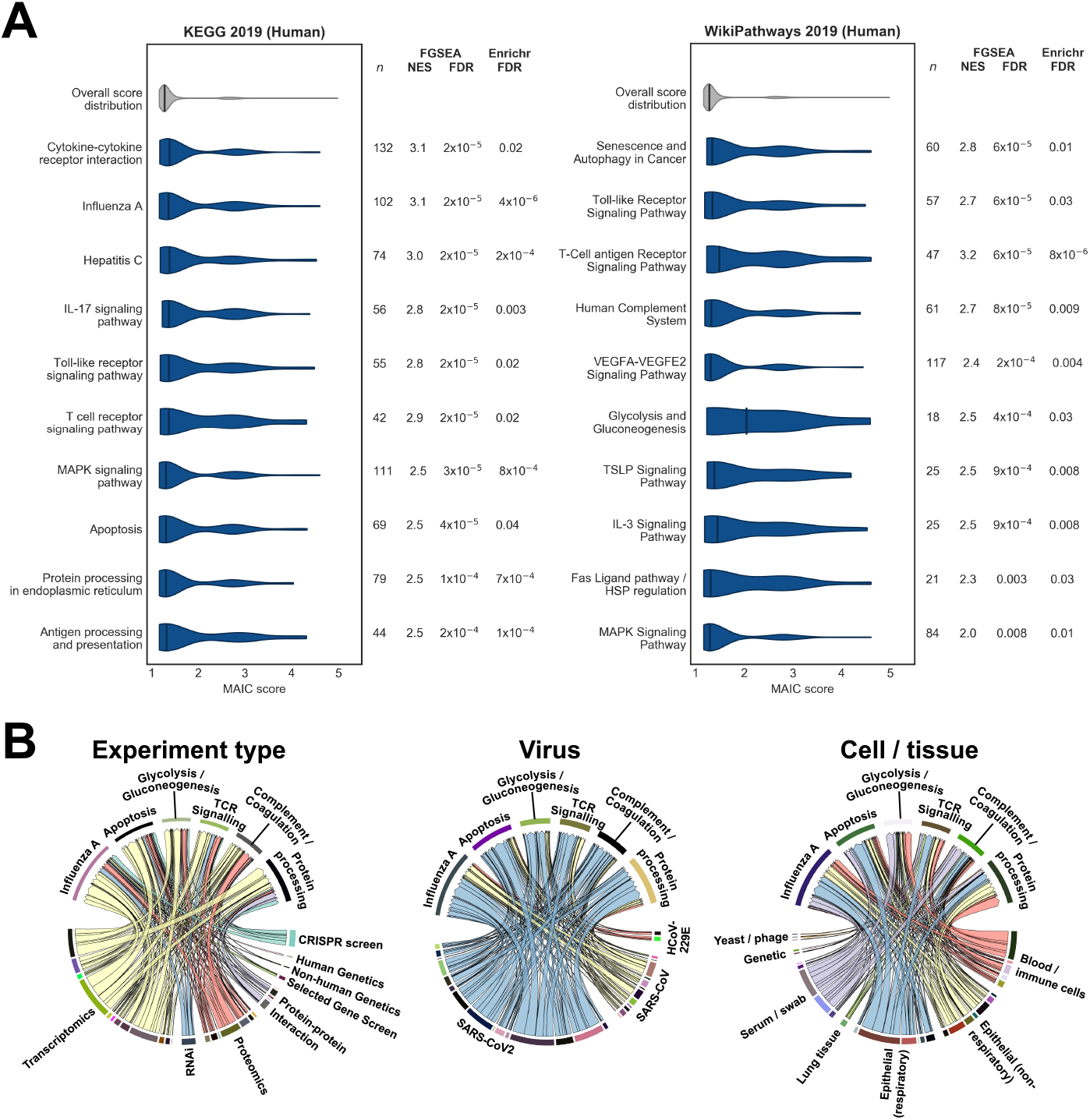
Gene Set Enrichment Analysis of MAIC rankings. (A) Violin plots of MAIC score distributions of top enriched pathways significant with both FGSEA and Enrichr algorithms, from the KEGG 2019 (Human) and WikiPathways 2019 (Human) databases. Highly similar pathways and irrelevant specific disease terms are not shown. *n*: number of gene set members included in the overall MAIC output; NES: normalised enrichment score from FGSEA. (B): Information contribution by methodology for selected enriched KEGG terms. Relative contributions of different information sources vary between functional annotations, but no single methodology predominates to drive enrichment.

Functional annotations that were significant using both methods and that were reflected in results from more than one database included terms related to cytokine, toll-like receptor and T-cell receptor signalling, protein processing, apop-tosis, the complement system, VEGF signalling, glucose metabolism and viral infections such as influenza A. As expected, the relative information contributions from different experiment types varied between pathways (fig. 4 B). For example, terms related to complement and coagulation received relatively little contribution from CRISPR screen data (derived from epithelial cells) but more information from proteomics and experiments using serum or swab samples, whilst pathways related to protein processing had relatively greater contributions from protein interaction studies. In all cases, enriched pathways drew information from a range of experiment types.

### Integration of MAIC output with results from published GWAS

One of the principal applications of MAIC is in the interpretation of results of genome-wide association studies (GWAS). Genome-wide association studies often implicate a locus containing a number of candidate genes and the precise nature of the interaction between gene and disease may not be known. As an example, we applied our results to the locus in chromosome 3 associated with hospitalisation due to COVID-19 in the sole COVID-19 GWAS published to date (data from which was also included in this analysis).^16^ This locus contained six genes (*SL6A20, LTZFL1, CCR9, FYCO1, CXCR6* and *CXR1*) that could all plausibly be linked to COVID-19 pathophysiology on the basis of their known functions.

Of these, *FYCO1*, which encodes a protein involved in vesicle transport and autophagy, was highly ranked in our results (rank 42). *FYCO1* is supported by SARS-CoV-2-specific protein-protein interaction and transcriptomic data. *CCR9* (rank 417) had additional support from a single transcriptomic study, while *SL6A20, LTZFL1, CXCR6* and *XCR1* had low ranks in our results, with no corroborating evidence in other studies.

## Discussion

The interpretation of any meta-analysis is critically dependent on the criteria for inclusion. In this case, our objective is to cast the net wide, including a range of data sources that are both conceptually and methodologically divergent. Experimental results bearing little relation to the composite of evidence from the other studies are downgraded by the MAIC algorithm, so the effect of irrelevant, noisy, or poorly-conducted experiments is minimal.^3^ By using permissive inclusion criteria, together with weighted meta-analysis, we have identifed key elements of the host-pathogen interaction and promising therapeutic targets for further investigation and intervention. These include host factors involved in viral replication, and elements of the immune response, which have been overlooked in the contributing studies (fig. 2).

### Key host factors related to viral entry and replication

Coronaviruses hijack host endomembranes to facilitate anchoring of the replication/transcription complex.^17^ Consistent with this, we observe an over-representaion of endoplasmic reticulum-related genes. Genes related to the function of the endoplasmic reticulum (*FAF2, ERGIC1, TENT5C, TENT5A, CFL1, STARD5*) and glycosylation (*MOGS, UGDH, UGGT1, PDIA6*, *PDIA3*) are observed along with a number of chaperone proteins (*HSPB1, HSPA4, HSPA8, HSP90AA1, ST13*). Many of these genes are related to the unfolded protein response (UPR), a stress response initiated by accumulation of mis-folded proteins. *FYCO1*, implicated in published GWAS results, has been suggested as a key mediator linking ER-derived double membrane vesicles, the primary replication site for coronaviruses, with the microtubule network.^18^

Viral entry via spike (S) protein-mediated membrane fusion is well characterised.^19^ The first step, spike activation, requires cleavage via host proteases such as cathepsin L *(CTSL*, rank 31).^20^ In the Wei *et* al.^21^ CRISPR screen, knockout of *CTSL* restricted viral production. Cathepsin L inhibition has been suggested as a promising therapeutic strategy for COVID-19: specific small molecule inhibitors are in early stages of development, and direct or indirect inhibition is observed with a number of approved drugs including glycopeptide antibiotics, chloroquine and dexamethasone.^22^ *ATP1A1* (rank 35), encoding a subunit of the NA+/K+ cotransporter, has similarly been shown to be necessary for membrane fusion and viral entry for a number of coronaviruses.^23^ Inhibition of *ATP1A1* by cardiac glycosides suppressed MERS-CoV infection in vitro.^24^ Additional anti-inflammatory^25^ effects of these drugs make them theoretically attractive therapeutic options, but adverse effects may limit their practical application.

The interferon-stimulated gene *LY6E* (rank 3), which plays a key role in enhancing cellular entry by RNA viruses including influenza A virus,^26^ was unexpectedly found to have a strong restricting effect on SARS-CoV-2, SARS-CoV and MERS-CoV.^27^ Such widely opposite differential effects on different viruses have been reported for other host genes, such as *IFITM3, RSAD2* and *AXL* (ranked 1056, 1039 and 215 respectively by MAIC),^26^ and for *PPIA* (see below).

### Immune response to SARS-CoV-2

Consistent with the emerging understanding of the pathogenesis of COVID-19, key genes in the inflammatory response to SARS-CoV-2 infection are highly represented in the top 100 genes. These include genes involved in recognizing the virus *(TLR3, IFIH1)*, activating the innate immune system *(OAS2, HERC5, S100A9)*, chemotaxis *(S100A9, CXCL10, CXCL8, CCL20, SAA2)* and pro-inflammatory cytokines *(IL1A, IL18)*. Toll-like receptor 3 *(TLR3)* is an endosome-associated pathogen-associated molecular pattern receptor, constitutively expressed in the respiratory tract and many immune cells. TLR3 detects double-standed viral RNA and triggers production of type I interferons and other pro-inflammatory cytokines, such as IL6 (rank 104) and TNFa (rank 182) via IRF3 and NF-*κ*B.^28^

The chemokine CXCL10 (rank 17) is a key signalling molecule in viral immunity, which could contribute to pulmonary inflammation as well as aiding viral clearance.^29^ CXCL10 levels are associated with outcome in influenza^30^ and are thought to have a protective effect in SARS, but a pro viral effect in HIV.^31^ CXCL10 has been proposed as a prognostic marker for the progression of disease in COVID-19, with continuously high levels of CXCL10 associated with worse outcomes.^32^

The high rankings of genes associated with activation and binding of T lymphocytes *(CD4, CD3E*, *FGL2*, *LCK, SELL)* are also likely related to their prognostic significance, as lymphopaenia is strongly associated with poor outcomes in COVID-19.^33,34^ Absolute counts of CD3+, CD4+ and CD8+ T lymphocytes have been proposed as a potential predictor of outcome in severe COVID-19 patients^35^ with an increase in numbers of these cells observed during recovery.

### Prioritisation of host susceptibility factors as therapeutic targets

The highest-ranking gene is *PPIA*, which encodes peptidyl-prolyl cis-trans isomerase A (*PPIA*, also known as cyclophilin A, *CypA*), a cytosolic protein involved in protein folding and trafficking, cell signalling and T-cell activation via the calcineurin/NFAT pathway.^36^ CypA is a pro-viral factor for hepatitis C virus (HCV), HIV-1, and SARS-CoV, and an anti-viral factor for IAV.^37,38^

The cyclophilin inhibitor cyclosporine has in vitro antiviral activity against HCV.^39,40,41^ This was also observed in a HCV clinical trial, where cyclosporine combined with interferon-a was more efficacious in achieving sustained virologic response than interferon monotherapy.^42^ Similar in vitro and clinical results were demonstrated for the CypA inhibitor alisporivir (DEBIO-015).^43^ CypA is also a pro-viral factor for HIV-1 and alisporivir can inhibit HIV-1 replication in vitro.^44,45^

A genome-wide protein-protein interaction screen identified an interaction between the SARS-CoV Nsp1 protein and CypA.^46^ Cyclosporine inhibits SARS-CoV replication in Vero E6 cells, as well as HCoV-229E, HCoV-NL63, avian coronavirus and feline coronavirus. Nsp1 also induced IL-2 expression in HEK293 cells through the calcineurin/NFAT pathway, making inhibition of this pathway interesting from both an antiviral and immunomodulatory perspective.^47^

The high ranking of *IL1A* (encoding interleukin *1-α*) is striking because monoclonal antibodies against interleukin 1 receptor are a plausible therapeutic target for COVID-19.^10^ This pro-inflammatory cytokine, which is synergistic with TNF*α*, is constitutively expressed in epithelial cells and is upregulated after SARS-CoV-2 infection.^48^ Interleukin-1 receptor blockade with anakinra is now being tested in a number of randomised clinical trials in COVID-19.^10^

### Advantages and limitations of data integration via MAIC

The principal advantage of the MAIC approach is that it allows integration of data from diverse sources. Unlike other methods for gene list comparison such as vote counting or robust rank aggregation,^49^ MAIC applies a data-driven weighting to each dataset, accepts both ranked and unranked lists, and includes user-defined categories which prevent any single method from overwhelming the results. MAIC outperforms other methods for predicting antiviral genes.^3^

This meta-analysis is restricted to studies involving genome-wide hypotheses or screening data for large gene sets, and does not consider evidence from candidate gene genetic studies or single-gene perturbations. Where a single gene has been investigated extensively but genome-scale studies are sparse, our approach may underestimate the relative strength of evidence for certain genes. Single gene studies, however, are likely to focus preferentially on genes that fit pre-conceived ideas of disease pathogenesis and may be prone to other biases such as publication bias, something which we mitigated against in our inclusion criteria.

Genetic perturbation data are still relatively sparse for SARS-CoV-2 and other human betacoronaviruses: only one genome-wide CRISPR knockout screen and two other sub-genome-scale screens (kinome-wide RNAi and interferon-stimulated gene overexpression screens) were included in the meta-analysis. Limited data of this type could be responsible for the lower than expected rankings for *ACE2* (rank 320), a major functional receptor for the SARS-CoV and SARS-CoV-2 spike (S) proteins, and *TMPRSS2* (rank 3037), a serine protease required for S protein priming.^19,50,51^ While *ACE2* was identified as a host dependency factor in the CRISPR screen, *TMPRSS2* was not, and as neither gene was included in the other two screens, the effects (or lack thereof) could not be confirmed. The only other supportive evidence for a role in disease pathophysiology, in studies included here, came from a single transcriptomic study for each; there was no evidence from protein-protein interaction, proteomics or genetics.^48,52^ Both of these genes have been proposed as possible therapeutic targets for COVID-19, and clinical trials are underway for the TMPRSS2 inhibitors nafamostat and camostat mesylate.

Systematic review and meta-analysis are routine elements in the assessment of clinical evidence and some fields in genomics, but have been less widely applied to mechanistic biology. Using a flexible and intuitive method, we have systematically reviewed and meta-analysed host gene-level data from studies that address a range of complementary questions regarding human betacoronavirus infection. This provides external validation for numerous host genes implicated in both viral life cycle, and immune response, and identifies several plausible therapeutic targets with broad support from multiple sources.

## Declarations

The authors declare no competing interests. All datasets included are publicly available or are available by reqest to the original authors.

## Data Availability

All data is available in the referenced papers or by contacting authors of the referenced papers.

https://baillielab.net/maic/covid19

## Acknowledgements

The authors would like to acknowledge Prof. C. Wiley and Prof J. Doench for generously sharing unpublished data and for their helpful comments on the manuscript. Work on this project was funded by UKRI (BBSRC and Medical Research Council [grant MC_PC_19059]).

## Notes

### Competing Interest Statement

The authors have declared no competing interest.

### Funding Statement

The authors would like to acknowledge funding from the following sources:
BBSRC, Wellcome Trust, MRC, Sepsis Research (FEAT) and the Intensive Care Society

### Summary of Updates

Updated title and abstract.

